# Exploring the Value of Peer Letters of Recommendation in the Holistic Review of Surgery Residency Applications: A Qualitative Study

**DOI:** 10.1101/2025.03.16.25323986

**Authors:** Caitlin Silvestri, Viemma Nwigwe, Subhash Krishnamoorthy, Cary B. Aarons

## Abstract

**Objective:** Residency applications rely on traditional letters of recommendation (*t*LORs) from faculty or mentors to evaluate applicants. However, interpretation of *t*LORs can be limited by potential biases, overuse of hyperbolic language, and a lack of longitudinal contact. We aimed to assess whether incorporating peer letters of recommendation (*p*LORs) would add a complementary perspective to the holistic review of an applicant’s attributes and potential.

**Design:** All applicants to a single, university-based general surgery residency program were invited to submit an *optional p*LOR in the 2023-24 recruitment cycle. Thematic analysis of applicants’ *p*LORs and *t*LORs was performed to identify patterns and sentiments.

**Setting:** Large general surgery residency program at a single, tertiary academic center.

**Participants:** Applicants selected for an interview for a general surgery residency program who submitted a *p*LOR in addition to their *t*LORs (*n*=95).

**Results:** Ninety-five applicants (78%) selected for interview submitted a *p*LOR along with their standard application to the categorical (*n=*77) and preliminary (*n=*18) tracks. Peer letter writers knew applicants for an average of 6.14 years (SD 4.7). Thematic analysis identified notable differences in *p*LORs: 1) peer letter writers more often evaluated applicants across diverse settings (professional and personal) over longer time periods, 2) *p*LORs placed greater emphasis on the applicants’ impact on others (peers, individuals, patients), and 3) provided more specific, tangible examples of each positive attribute. Lastly, *p*LORs summative assessments often included personal language while *t*LORs tended to stratify applicants using percentiles or coded language.

**Conclusion:** Peer letters of recommendation offer a unique, complementary perspective in the holistic residency application review process. Compared with traditional letters, *p*LORs provide a richer context of an applicant’s impact in a community of their peers, more often providing tangible examples. This perspective is crucial for evaluating applicants as we build diverse and collaborative learning communities each year.

**Funding:** This research did not receive any specific grant from funding agencies in the public, commercial, or not-for-profit sectors.

**ACGME Core Competencies:** Professionalism (P), Interpersonal and Communication Skills (ICS)

**Highlights:**

- Peer letters of recommendation (*p*LORs) offer a unique perspective in applicant review
- *p*LOR writers assess applicants across diverse settings over longer time periods
- *p*LORs emphasize applicants’ community impact with tangible, specific examples
- *t*LORs use ranking language, while *p*LORs include more personal, narrative assessments
- pLORs complement *t*LORs in holistic residency selection and evaluation processes

## Introduction

When applying for general surgery residency, medical student applicants submit both subjective and objective data through the Electronic Residency Application Service (ERAS)^1^, including but not limited to a personal statement, curriculum vitae (CV) components, medical school transcripts, United States Medical Licensing Exam (USMLE) scores, and letters of recommendation (LORs)^2^. Historically, general surgery residencies review and place emphasis on an applicant’s academic metrics such as exam scores, clerkship grades, publications, research experience, and Alpha Omega Alpha (AOA) membership^3–5^. However, these quantitative measures of evaluation do not necessarily capture the full breadth of an applicant’s potential, and studies have demonstrated variability in their ability to predict success in residency^6,7^.

Beyond academic metrics, residency selection committees also consider qualitative components such as personal statements and LORs. The role of the personal statement in the recruitment process remains inconsistent, with program directors differing in how they use it for interview selection and ranking^8^. Furthermore, studies have revealed substantial interreader variability, with a single personal statement often receiving multiple conflicting evaluations^9,10^. Similarly, LORs are a required element of the ERAS application and are intended to provide insight into an applicant’s abilities, accomplishments, and readiness for residency. Traditional letters of recommendation (*t*LORs), typically authored by department chairs, faculty, or mentors, offer insights into an applicant’s performance based on personal interactions and peer comparisons.

However, their reliability and impact in the selection process have been increasingly scrutinized due to concerns about bias, high interreader variability, and inconsistent influence on decision-making^11–14^. Additionally, LORs are frequently overly laudatory, contain hyperbolic language, and rely on coded phrasing that can obscure meaningful distinctions between applicants, further complicating their role in the selection process^15,16^.

Recognizing the limitations of our current evaluation processes and metrics, the Association of American Medical Colleges (AAMC) developed a holistic review framework, which seeks to assess applicants as whole individuals rather than relying solely on academic achievements ^17^. Holistic review allows for flexible, individualized, and mission-driven applicant review, aligning with the growing focus on improving diversity and equity in the physician workforce. Efforts to enhance holistic review have led to notable changes in the residency application process. One of the most significant shifts was the transition of the USMLE Step 1 to a pass/fail scoring system by the National Board of Medical Examiners (NBME), a move aimed at encouraging a broader evaluation of applicants^18^. This transition has created an opportunity for programs to redefine selection criteria and prioritize other factors, including interpersonal skills, leadership, and professionalism.

As residency selection continues to evaluate candidates, LORs remain a key component of holistic review, despite their well-documented limitations. To provide a more comprehensive evaluation of applicants, new strategies should be explored to supplement *t*LORs. One potential tool is the inclusion of *p*LORs, which unlike *t*LORs, are authored by peers such as classmates, co-workers, friends, teammates, or members of shared organizations who can also offer unique perspectives on an applicant’s qualities. *p*LORs have been utilized in the undergraduate school admission process ^19^, but have not yet been used in the residency recruitment.

The introduction of *p*LORs in residency recruitment may help bridge the gaps posed by *t*LORs and other traditional application metrics by providing a different, complementary perspective of an applicant’s potential. The primary objective of this study is to evaluate the contextual differences between *t*LORs and *p*LORs for applicants to a single university-based general surgery residency program. We hypothesize that *p*LORs will offer a richer, complementary perspective compared to traditional faculty-authored letters, ultimately strengthening the holistic review process.

## Methods

This study was reviewed and deemed exempt from full review by the Columbia University Institutional Review Board (IRB) under protocol number AAAU9012. In accordance with ethical research guidelines, participant confidentiality was maintained throughout data collection and analysis. No identifiable applicant information was included in the study. This study adheres to the Standards for Reporting Qualitative Research (SRQR)^20^ to ensure transparency, rigor, and completeness in reporting.

### Participants

A single, university-based general surgery residency program invited all applicants to submit an optional *p*LOR during the 2023-24 residency recruitment cycle. The study included applicants who received an interview invitation and submitted both an optional *p*LOR and at least one *t*LOR. No applicants were excluded based on demographics, specialty interests, or letter content. After the residency match process concluded, we retrospectively reviewed the data, ensuring that our analyses did not influence recruitment or selection outcomes.

### Data Collection

The Electronic Residency Application Service (ERAS), managed by the Association of American Medical Colleges (AAMC)^1^, collects *t*LORs as part of the standard residency application process. Faculty members and department chairs typically write these letters. To ensure consistency, we selected a single *t*LOR per applicant, specifically the first *t*LOR written by a surgical faculty member within the ERAS application packet.

Applicants submitted a single, optional, *p*LOR separately from their ERAS application. Each applicant identified a peer—such as a close friend, classmate, co-worker, teammate, research collaborator, or member of the same interest group or organization—to write a recommendation. Applicants were explicitly instructed that the letter could not be written by a family member, someone in a supervisory role, or a classmate applying into the same specialty. Peer letter writers were instructed to submit their letters directly to the residency program, with approximately 10 days to complete them from the time the application was downloaded by the program. When submitting the *p*LOR, letter writers also provided demographic information, including the nature of the relationship to the applicant and length of time known.

### Reflexivity

Author [BLINDED-C], a male Professor of Surgery, serves as the Program Director (PD) of a general surgery residency program and Vice Chair of Education (VCE) in the Department of Surgery. He holds a medical degree and a Master’s of Science in Education (MSEd), where he received formal training in qualitative research methods. Before initiating this study, he participated in the residency application process as program director, overseeing the holistic review process for interview selection and conducting applicant interviews. His perspective provided a broader contextual understanding that informed the thematic analysis.

Authors [BLINDED-A] and [BLINDED-B], female general surgery residents, collaborated with [BLINDED-C] in data coding and analysis. Both had received informal training in qualitative research methods and had prior experience in the residency application review process, contributing to holistic applicant evaluations for interview selection. Unlike [BLINDED-C], they did not conduct applicant interviews during the study cycle, minimizing potential bias in the generation and interpretation of themes.

### Analytic Approach

Author [BLINDED-C] de-identified all LORs by removing identifiable information, including medical school names, applicant names, and AAMC ID numbers, before analysis. The authors used Taguette (version 1.4.1-60-ga73a5e2), a free qualitative analysis software, to organize coding and develop codebooks.

Using an open, inductive coding approach, authors [BLINDED-C], [BLINDED-A], and [BLINDED-B] independently reviewed all *p*LORs and *t*LORs and developed separate codebooks based on emergent content. Throughout the coding process, the authors conducted debriefing sessions to refine initial codes and definitions, ensuring alignment and consistency. After finalizing codebooks, each author re-coded their assigned LORs to reflect the refined framework.

Following individual coding, all authors collaboratively constructed themes through group discussions and consensus-building. Due to the de-identified nature of the data, we could not conduct member checking by inviting participants to review the identified themes. However, we ensured trustworthiness and rigor through investigator triangulation via independent coding by multiple authors, peer debriefing sessions, and consensus-based construction of final themes^21^.

## Results

### Participant Demographics

During the 2023-2024 recruitment period, the program received approximately 1600 applications to the general surgery residency, including both categorical and preliminary tracks. Of these, 122 applicants were selected for interviews, including 83 (68%) for the categorical track and 39 (32%) for the preliminary track. Among interviewees from both tracks, 95 (78%) submitted an optional *p*LOR along with their standard ERAS application. Of the 95 applicants included in the analysis, 77 (81%) were categorical and 18 (19%) were preliminary track applicants (Table 1).

**Table 1.** Applicant demographics of the general interview pool vs. *p*LOR submission pool

| Characteristic | Total Interview Pool<br>Frequency (%),<br>N=122 | *p*LOR Submissions<br>Pool Frequency (%),<br>N=95 |
| --- | --- | --- |
| Gender |  |  |
| Female | 72 (59%) | 61 (64%) |
| Male | 50 (41%) | 34 (36%) |
| Application Track |  |  |
| Categorical | 83 (68%) | 77 (81%) |
| Preliminary | 39 (32%) | 18 (19%) |

### pLOR Writer Demographics

Among the 95 submitted *p*LORs, the writers had known the applicants for an average of

6.14 years (SD 4.7, range 0.2–23 years). The primary relationship between the letter writer and the applicant varied, with the most common being friends (*n*=55, 58%) and classmates (*n*=32, 34%). Less common relationships included teammates (*n*=3, 3%), research collaborators (*n*=2, 2%), co-workers (n=2, 2%), and coaches (*n*=1, 1%) (Table 2). Additionally, 15 (16%) of *p*LOR writers indicated they had multiple relationships with the applicant.

**Table 2.** Primary relationship of *p*LOR writers to the applicant

| Characteristic | Total Interview Pool<br>Frequency (%), N=95 |
| --- | --- |
| Friend | 55 (58%) |
| Classmate<br>(*not applying in same specialty*) | 32 (34%) |
| Teammate | 3 (3%) |
| Research Collaborator | 2 (2%) |
| Co-worker | 2 (2%) |
| Coach | 1 (1%) |

### Overarching Themes

We compared *p*LOR and *t*LOR submissions and identified four major themes (Table 3).

**Table 3.** Thematic analysis of *p*LORs and *t*LORs

| Theme | Description | Example |
| --- | --- | --- |
| **Qualifications** | ***tLORs*** - evaluators impart their appointments & credentials to support their qualifications to assess the applicant and other students.<br>***pLORs*** - evaluators establish the length of time & various contexts in which they know the applicant as evidence of their qualifications to make a recommendation. | ***tLOR:*** "As director of our student education programs and of the Surgical Education Fellowship, I have had close and direct involvement in observing, assessing and interacting with our students. (P14)"<br>***pLOR:*** "I have known [APPLICANT] for three years as a classmate and close friend. In this capacity, I can comment on his personal characteristics as well as academic engagements. (P15)" |
| **Positive Attributes** | ***tLORs*** – applicant attributes described are focused primarily on clinical performance, technical skills, & cognitive ability. More often includes input from others (joint letters, imported quotations).<br>***pLORs*** – applicant attributes described are more focused on | ***tLOR:*** "He seemed to thrive in that environment and impressed us all with his clinical, technical acumen, his rapid progress and his extraordinary work ethic and learning efforts." (P14)<br>***pLOR:*** "One specific example was during a group project we worked on together in our neurobiology class. Our task was to analyze a complex case study and present our findings to the class. [APPLICANT] took charge of coordinating the project and immediately led a |
|  | relationships & character with less emphasis on clinical & technical skills. Specific, supporting examples are frequently provided. | *weekly discussion group with other students. [APPLICANT] also encouraged open communication and brainstorming, creating a safe environment where everyone felt comfortable sharing their thoughts and suggestions.” (P58)* |
| **Roles and Accomplishments** | **tLORs** - evaluators typically recapitulate awards, grades, USMLE scores, leadership roles, research, and publications that can be found on an applicant’s CV.<br>**pLORs** - evaluators often discuss roles and accomplishments within the context of a community. The focus is on the applicant’s impact on individuals, groups, & patients. | **tLOR:** “After coming to [Institution], his scholarly activity continued to be outstanding, with Alpha Omega Alpha (AOA) recognition, many Honors grades, a notable USMLE Step 2 CK score of 275, and numerous research activities.(P24)”<br>**pLOR:** “[APPLICANT] is well known amongst her peers as a go-to person for many medical school related concerns. I have personally witnessed [APPLICANT] mentor and guide many students through the early years of their medical training. Peers regularly refer students to [applicant] when they need a listening ear, resources, or advice navigating their way through all steps of the medical school journey.” (P8) |
| **Summative Assessment** | **tLORs** - attempt to stratify applicants using percentiles & coded language. Expresses a desire to keep students in their programs (if applicable).<br>**pLORs** - summative assessments and recommendations are often more personal. Expresses a desire for applicants to take care of their family or work with them as a colleague. | **tLOR:** “In summary, [APPLICANT] is easily within the top 1% of medical students I’ve had the honor of training... I will be heavily recruiting him for our own program.” (P16)<br>**pLOR:** “[APPLICANT] is going to be a phenomenal co-resident, and I truly envy his future colleagues. He is authentically kind, a team player, a hard worker, and a mentor to the learners around him. [APPLICANT] is the kind of person who I want treating my family members in the future. I give him my highest recommendation possible.” (P16) |

#### Qualifications

Both *p*LOR and *t*LOR writers commonly discussed their qualifications for writing the LOR, but they differed in the type of content used to establish their credibility to the reader.

*t*LOR writers typically emphasized their professional and academic roles, citing institutional appointments and leadership positions such as medical student clerkship director, department chair, or program director to support their ability to assess the applicant and compare to other students: “*As director of our student education programs and of the Surgical Education Fellowship, I have had close and direct involvement in observing, assessing and interacting with our students” (P14)*.

In contrast, *p*LOR writers more frequently established their qualifications by referencing the length of time and the variety of contexts in which they knew the applicant, positioning their personal experiences as foundations for their assessment of the applicant: *I have known [APPLICANT] for three years as a classmate and close friend. In this capacity, I can comment on his personal characteristics as well as academic engagements (P15)*.

#### Positive attributes

Both *p*LOR and *t*LOR writers consistently highlighted the applicant’s positive attributes throughout their letters, but they differed in the types of attributes emphasized and the ways in which they substantiated these qualities.

In *t*LORs, writers most commonly described attributes related to clinical performance, technical skills, and foundational medical knowledge. *“He seemed to thrive in that environment and impressed us all with his clinical, technical acumen, his rapid progress, and his extraordinary work ethic and learning efforts” (P14)*. These letters frequently incorporated input from multiple evaluators, often through composite letters, feedback from additional evaluators, or imported quotations from summative assessments: *“In preparation of this letter, I sought the opinion of the chief resident who worked with her as a sub-intern, which generated the following comments: “[APPLICANT] learns quickly, has a solid fund of knowledge and reasons well from first principles. She has a good attitude and is always willing to help the team. She routinely came in early and stayed late to make sure her patients were given optimal care. She was very helpful with floor work and quickly learned how to do routine patient care tasks. Her technical skills in the OR were very good for her level, certainly more advanced than I would expect for a medical student. She is very self-aware of any deficiencies and is always working hard to improve*.*” (P54)*.

In contrast, *p*LOR writers more frequently emphasized interpersonal relationships and character traits, with significantly less focus on clinical or technical abilities. Furthermore, *p*LOR writers tended to support these attributes with detailed, anecdotal examples illustrating the applicant’s behavior in specific contexts: *“One specific example was during a group project we worked on together in our neurobiology class. Our task was to analyze a complex case study and present our findings to the class. [APPLICANT] took charge of coordinating the project and immediately led a weekly discussion group with other students. [APPLICANT] also encouraged open communication and brainstorming, creating a safe environment where everyone felt comfortable sharing their thoughts and suggestions” (P58*).

#### Roles and accomplishments

Both *t*LOR and *p*LOR writers consistently highlighted the applicant’s roles and accomplishments, but they differed in the types of achievements they emphasized.

*t*LOR writers primarily focused on accomplishments typically found in the applicant’s CV, such as awards, grades, USMLE scores, leadership positions, research achievements, and scholarly contributions (e.g., publications, abstracts, and oral presentations). These letters often framed the applicant’s accomplishments within a structured, academic context: *“After coming to [Institution], his scholarly activity continued to be outstanding, with Alpha Omega Alpha (AOA) recognition, many Honors grades, a notable USMLE Step 2 CK score of 275, and numerous research activities” (P24)*.

In contrast, *p*LOR writers tended to discuss roles and accomplishments within the context of the applicant’s impact on their community—whether among peers, in research settings, or with patients. These letters highlighted the applicant’s influence on individuals and groups, often illustrating their mentorship, leadership, and support for others: *“[APPLICANT] is well known among her peers as a go-to person for many medical school-related concerns. I have personally witnessed [APPLICANT] mentor and guide many students through the early years of their medical training. Peers regularly refer students to [APPLICANT] when they need a listening ear, resources, or advice navigating their way through all steps of the medical school journey” (P8*).

#### Summative assessment

Both *t*LOR and *p*LOR writers included a summative assessment in the letter, but differed in how they characterized the applicant’s overall impact.

*t*LOR writers frequently used stratification methods, such as percentiles or coded language (e.g., outstanding, excellent, exceptional), to rank applicants relative to their peers. Additionally, they often indicated whether their institution would actively recruit the applicant to their program: *“In summary, [APPLICANT] is easily within the top 1% of medical students I’ve had the honor of training*… *I will be heavily recruiting him for our own program” (P16*).

In contrast, *p*LOR writers tended to offer summative assessments that were more personal, often expressing a desire to work with the applicant as a future colleague or entrust them with the care of loved ones and family members: *“[APPLICANT] is going to be a phenomenal co-resident, and I truly envy his future colleagues. He is authentically kind, a team player, a hard worker, and a mentor to the learners around him. [APPLICANT] is the kind of person I want treating my family members in the future. I give him my highest recommendation possible” (P16*).

## Discussion

This qualitative analysis compared traditional letters of recommendation (*t*LORs) and peer letters of recommendation (*p*LORs) for applicants interviewing at a single general surgery residency program. To our knowledge, our study is the first to incorporate *p*LORs into the residency recruitment process and qualitatively compare *t*LORs and *p*LORs. We identified four key themes illustrating how *t*LORs and *p*LORs capture complementary perspectives on applicants, albeit through different lenses. Specifically, *t*LORs tend to emphasize academic achievements, technical skills, and summative comparisons by employing coded language, percentiles, and a desire to recruit them. Compared to *p*LORs which provide a more personal appraisal by highlighting an applicant’s character, interpersonal relationships, engagement within the community, and the direct impact they have on peers and patients, all while providing more detailed examples of these traits.

Traditional LORs are a longstanding component of residency applications, but come with notable limitations. Up to 20% of *t*LORs are written by department chairs - close to half of whom have limited direct clinical experience with the applicant - or are written from faculty with single, narrow contact with the applicant^22^. Prior research has also shown that nearly 17% of *t*LORs lack evidence of direct clinical supervision in addition to being overly laudatory, and often including hyperbolic, coded, and biased language^22 11,23^. While key components of an applicants’ CV may become diluted within the ERAS application, *t*LOR writers frequently recapitulate what they find to be most important aspects including exam scores, clinical grades, awards, and research productivity. Lastly, we found that *t*LOR writers often employ summative language, including percentiles and coded language and frequently include whether the applicant will be recruited to their program - a factor that previous studies have identified as highly impactful for faculty in selection^24^. Despite their limitations, *t*LORs can still provide critical components within the residency recruitment process, offering programs valuable insights into attributes deemed essential for success in surgical training.

In an effort to address the various limitations of *t*LORs, some specialties have adopted standardized letters of recommendations (SLORs), which are intended to offer a more uniform, nationalized scale for evaluating applicants^25^. Surgical specialties have tried to implement SLORs, however despite their aim for consistency and applicant stratification, are still found to be overly favorable to applicants^25–28^. A SLOR was created for general surgery by the American College of Surgeons (ACS) and Association for Program Directors in Surgery (APDS)^29^, however, widespread adoption and utilization has not occurred. Focus groups with key stakeholders, including general surgery clerkship and program directors, have expressed reservations about SLORs due to performance inflation and reliance on scaled assessments of attributes without providing examples^23^. This further emphasizes the need for alternative approaches.

Our study demonstrates that *p*LORs can address some of these limitations by providing a complementary perspective of applicants. Unlike *t*LORs, *p*LOR writers generally have longer and more varied relationships with the applicants. Additionally, they focus on more personal attributes - such as character, interpersonal relationships, and community engagement - that are typically not captured in *t*LORs. *p*LORs also offer specific examples and narratives about an applicant’s impact on peers, their community, and patients - a context that is less common to read about it *t*LORs, but critical as we endeavor to create effective and diverse learning teams each year. Lastly, their summative assessment is a more personal reflection, discussing genuine desires to work with the applicant or having them care for a family member, rather than *t*LORs using percentiles and coded language to compare to their peers. This personal lens is particularly important in an era when objective data within residency recruitment is declining, as it provides program directors with a more nuanced and holistic review of an applicant’s potential.

This study has several key limitations. First, it was conducted at a single academic center and only evaluated LORs from applicants that had been selected for interviews, which may limit the generalizability. Moreover, the optional nature of the *p*LOR submissions could have introduced selection bias, with applicants who were able to secure a supportive peer recommendation might not represent the entire applicant pool, potentially skewing the overall portrayal more positively. Although our sample size was sufficient to reach thematic saturation, further research with larger and more diverse cohorts is necessary^30^.

Given these findings, we recommend that residency program directors consider incorporating *p*LORs as an optional component in the ERAS application. Such integration would allow programs that value perspectives on interpersonal relationships, communication, community impact, and applicants’ character to enhance their holistic review process. Future research should involve multi-institutional studies to validate our findings and explore the utility of *p*LORs in other specialties.

## Conclusion

In conclusion, our study demonstrates that *p*LORs provide valuable, complementary insights to traditional evaluations by emphasizing personal attributes and community impact that are often overlooked. Despite limitations such as single-center data and potential selection bias, these findings support the integration of *p*LORs into the residency recruitment process to enhance holistic review. Future multi-institutional studies will be essential to further validate and refine the use of *p*LORs in applicant assessment.

## Data Availability

All data produced in the present study are available upon reasonable request to the authors

## Abbreviations

*p*LORs: peer letters of recommendation
*t*LORs: traditional letters of recommendation

